# Online respondent-driven detection for enhanced contact tracing of close-contact infectious diseases: benefits and barriers for public health practice

**DOI:** 10.1101/2020.06.24.20138024

**Authors:** Yannick B. Helms, Nora Hamdiui, Renske Eilers, Christian Hoebe, Nicole Dukers-Muijrers, Hans van den Kerkhof, Aura Timen, Mart L. Stein

## Abstract

**Background:** Online respondent-driven detection (online-RDD) is a novel method of case-finding that can enhance contact tracing. However, the advantages and challenges of online-RDD for contact tracing (CT) have not yet been investigated from the perspective of public health professionals (PHPs). Therefore, it remains unclear if, and under what circumstances, PHPs are willing to apply online-RDD for contact tracing.

**Methods:** First, between March and April 2019, we conducted semi-structured interviews with Dutch PHPs responsible for CT in practice. Questions were derived from the ‘diffusion of innovations’ theory. Second, between May and June 2019 we distributed an online questionnaire to 260 Dutch PHPs to quantify the main qualitative findings. Using hypothetical scenarios that involved close-contact pathogens (scabies, shigella, and mumps), we assessed anticipated advantages and challenges of online-RDD and PHPs’ intention to apply online-RDD for contact tracing.

**Results:** Twelve interviews were held and 70 PHPs filled in the online questionnaire. A majority of questionnaire respondents (71%) had a positive intention towards using online-RDD for contact tracing. Anticipated advantages of online-RDD were related to accommodating easy and autonomous participation in contact tracing of patients and contact persons, and reaching contact persons more efficiently. Anticipated challenges with online-RDD were related to limited opportunities for PHPs to support, motivate, and coordinate the execution of contact tracing, adequately conveying measures to patients and contact persons, and anticipated unrest among patients and contact persons. Online-RDD was considered more applicable when patients and their contact persons are reluctant to share sensitive information directly with PHPs, digitally skilled and literate persons are involved, and the scope of contact tracing is large. Online-RDD was considered less applicable when consequences of missing information or individuals are severe for individuals - or public health, when measures that patients and contact persons need to undertake are complex or impactful, and when a disease is perceived as particularly severe or sensitive by patients and their contact persons.

**Conclusions:** PHPs generally perceived online-RDD as beneficial to public health practice. The method can help overcome challenges present in regular CT and could be used during outbreaks of infectious diseases that spread via close-contact, such as the SARS-CoV-2 virus. We propose a staggered implementation study to further investigate the application of online-RDD for enhanced CT during the ongoing COVID-19 pandemic.

## Introduction

Contact tracing (CT) is a pivotal control measure in the fight against infectious diseases that transmit through contact between humans, such as measles, tuberculosis, and SARS-CoV-2 (1–3). In practice CT meets certain challenges; most notably the timeliness of case-finding and the heavy workload associated with the CT process for public health professionals (PHPs) (4–6).

The current COVID-19 pandemic highlights some of these challenges. CT-efforts worldwide have been overwhelmed by the scope and intensity of CT required to keep up with the widespread and rapid transmission of the virus. As such, now more than ever, (technological) innovations that support and enhance CT are warranted, so that control of COVID-19, and other future outbreaks, may become more feasible for PHPs (7).

Several approaches have been proposed in this regard, in particular the use of mobile phone apps that measure and record proximity to others, and/or tracking patients’ mobile GPS-location history (3,7–9). Though promising, these approaches currently lack an empirical evidence base, have not yet been applied in practice, or may be sensitive from ethical, legal, or privacy perspectives (10).

Recently, a new method for case finding that is based on the principles of respondent-driven sampling was introduced that can enhance CT: online Respondent-Driven Detection (online-RDD) (11,12). Online-RDD can be used for the detection of cases or clusters of cases within contact networks. It starts with an index-case, who is asked to recruit contact persons (i.e. individuals that had contact with the index through which transmission of a pathogen might have occurred) through online means, such as through email, WhatsApp, or other online messaging platforms. Contact persons then fill out a web-based questionnaire on disease symptoms and/or pre-specified relevant behaviours (by exposure risk, e.g. visited locations) to assess the risk of, or identify, a potential infection. If necessary, contact persons are asked to 1) contact a PHP for consultation and/or testing, and 2) recruit new contact persons to do the same, and so on (see Fig 1 for a schematic overview of online-RDD for CT). With direct (peer-to-peer) and online communications, online-RDD potentially accelerates CT and lowers the workload for PHPs.

**Fig 1.**
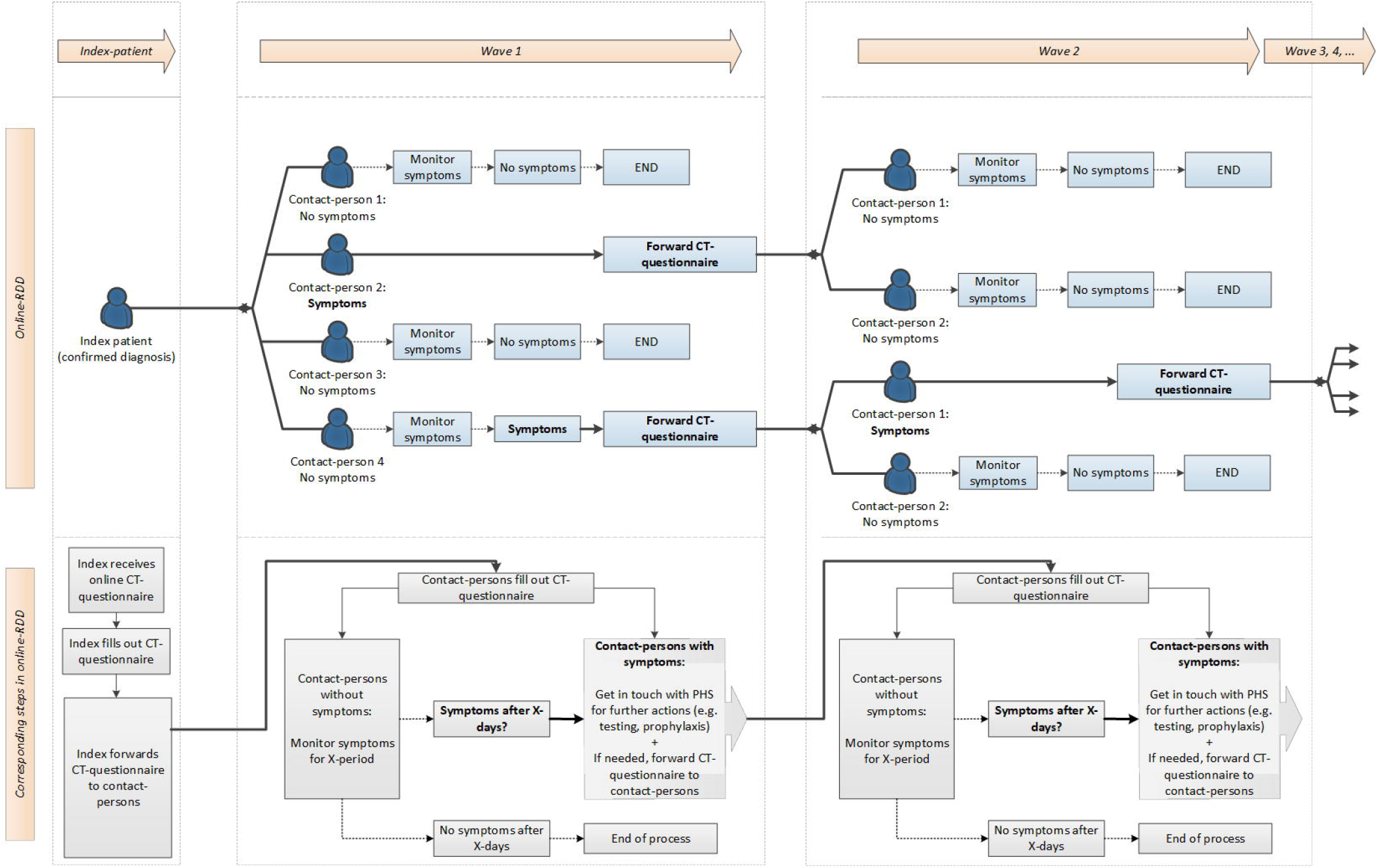
Online-RDD for CT schematic overview.

So far, online-RDD has been used in research settings (12–14), and logistics were tested in practice in a preparatory small scale pilot studies on measles and pertussis in the Netherlands (unpublished results). However, (inter)nationally the potential application of online-RDD for CT has not been systematically investigated from the perspective of PHPs involved in CT. As such, to understand the implementation potential of online-RDD for CT, a needs assessment in practice is urgently needed. Therefore, the aims of this study were to investigate what advantages and challenges PHPs anticipate with regard to the use of online-RDD for CT in practice, under what circumstances online-RDD can be used for CT, and how they would consider its application in practice. In this study, we focused on infectious diseases that spread through close contact between people, namely scabies, mumps and shigella. As such, we believe that our results are especially relevant in the current context of COVID-19. Therefore, we conclude by suggesting steps towards implementation of online-RDD for CT during the current COVID-19 pandemic.

## Materials and methods

We conducted a sequential exploratory mixed methods study (15). First, qualitative data were collected and analysed (Phase 1). Subsequently, these data were used to develop an online questionnaire, through which we quantified our qualitative findings (Phase 2).

### Phase 1: qualitative data collection and analysis

#### Sampling

In the Netherlands, infectious disease control is executed by PHPs (nurses and doctors) working for public health services (PHS). There are 25 PHS in the Netherlands, which serve different socio-geographical regions. We selected three PHS from different regios (Utrecht, South-Holland, and South-Limburg) and asked them to invite two or three nurses and doctors with experience in CT for close-contact pathogens to partake in our interviews.

#### Data collection and development of research materials

Semi-structured interviews with PHPs were conducted in Dutch by one male interviewer (YBH) from March through April 2019. Interviews were held at interviewees’ respective PHS, and lasted approximately one hour. We developed an interview guide based on the ‘innovation decision-process’ model from Rogers’ ‘diffusion of innovations’ (DOI) theory (16). Following this model, a PHPs’ intention to adopt (to make use of) online-RDD for CT is influenced by 1) prior conditions, such as existing CT-practices, 2) user characteristics, such as PHPs’ age and work experience, 3) anticipated attributes (characteristics) of online-RDD, such as its relative advantage, compatibility, complexity, observability, and trialability, and 4) communication channels through which online-RDD is communicated to PHPs. For the purposes of this study, we focused on topics 1 through 3. Topic 4 is part of the next step, which can be found as our recommendations for the implementation-process.

##### Interviews consisted of four parts

First, exploratory questions regarding PHPs’ experiences with CT were asked, focusing on their perceptions of current CT practices.

Second, since at the time of the interviews most PHPs were unfamiliar with online-RDD, we introduced online-RDD to interviewees using a PowerPoint presentation (based on Fig 1), designed for this purpose. A standardised script for the explanation of online-RDD was used to ensure all interviewees received the same information.

Third, the potential application of online-RDD for CT was discussed with interviewees in the context of three hypothetical scenarios. This is an effective method to elicit perceptions and attitudes towards certain actions in a real-life context (17). To develop realistic and relatable scenarios, we developed these in collaboration with PHPs working at the Dutch National Coordination Centre for Communicable Disease Control, which is part of the National Institute for Public Health and the Environment (RIVM), in the Netherlands.

Scenarios used in this research differed in terms of the disease and respective guidelines for CT, the index-patient’s background (work and living situation), and the contact persons potentially involved (for a detailed description of the scenarios, see S1 Figs):

- Scenario 1, ‘Scabies’: A student living in a student housing complex, who was diagnosed with scabies after having had experienced symptoms for approximately three months.
- Scenario 2, ‘Shigella’: A middle-aged individual who was diagnosed with shigella upon returning to his home country from an organised group holiday with friends.
- Scenario 3, ‘Mumps’: A student with a side-job as baby-sitter, who was diagnosed with mumps.

For each scenario, interviewees were asked whether and how they would consider applying online-RDD for CT and why (not), and what they considered advantages and challenges of this approach. Thereafter, interviewees were asked to rank the scenarios in terms of their suitability for applying online-RDD, and to explain their ranking.

Fourth, the potential application of online-RDD for CT in interviewees’ ‘day-to-day practice’ was discussed.

The interview guide and the materials used during interviews (PowerPoint introduction to online-RDD and scenarios) were extensively pilot tested with a small sample of PHPs at the Dutch National Coordination Centre for Communicable Disease Control. Interviewees were verbally informed about the study and asked for their written consent before each interview. Each participant was given a small (non-monetary) token of appreciation after the interview.

#### Data analysis

All interviews were audio-recorded and transcribed ad verbatim. We conducted a thematic analysis (18) to identify advantages and challenges of online-RDD for CT, and characteristics of scenarios that enable or restrain the use of online-RDD for CT. Open-, axial-, and selective coding were conducted in MAXQDA 2018 v.18.0.5. No new (sub) themes emerged (i.e. data saturation was reached) after eight interviews. Nevertheless, we conducted four more interviews, as these were planned with a different PHS. Of all interviews, 25% was randomly selected to be double coded by a second researcher (NH). Divergent findings were discussed until consensus was reached.

### Phase 2: quantitative data collection and analysis

#### Sampling

Two-hundred and sixty PHPs from all 25 PHS in the Netherlands were invited via email to complete an online questionnaire between May and June 2019. Questionnaires were accessible to respondents for four weeks, during which we sent three reminders.

#### Online questionnaire

An online questionnaire was developed to quantify our main qualitative findings from Phase 1. To this end, we formulated statements regarding the qualitatively identified advantages and challenges of online-RDD for CT, and regarding intention to use online-RDD for CT (S2 Table 1). Questionnaire respondents could respond on a 5-point Likert scale, ranging from strongly agree (1) to strongly disagree (5). The online questionnaire contained four sections:

**Table 1.** Interviewee’s & questionnaire respondents’ characteristics.

|  | Interviewees (n=12) | Respondents (n=70) |
| --- | --- | --- |
| **Age, in years (M;IQR)** | 38.5 (34.5-56.8) | 49 (36-59.3) |
| **Sex (%)** |  |  |
| - Male | 4 (33.3) | 22 (31.4) |
| - Female | 8 (66.7) | 48 (68.6) |
| **Province of employment (%)** |  |  |
| - Brabant | . | 6 (8.6) |
| - Caribbean Netherlands* | . | 1 (1.4) |
| - Drenthe | . | 1 (1.4) |
| - Flevoland | . | 1 (1.4) |
| - Friesland | . | 2 (2.9) |
| - Gelderland | . | 14 (20.0) |
| - Groningen | . | 5 (7.1) |
| - Limburg | 4 (33.3) | 7 (10.0) |
| - North-Holland | . | 13 (18.6) |
| - Overijssel | . | 6 (8.6) |
| - Utrecht | 5 (41.7) | 4 (5.7) |
| - Zeeland | . | 1 (1.4) |
| - South-Holland | 3 (25) | 9 (12.9) |
| **Role (%)** |  |  |
| - PHS-nurse | 6 (50.0) | 33 (47.1) |
| - PHS-doctor | 6 (50.0) | 35 (50.0) |
| - PHS-managers | . | 2 (2.9) |
| **Experience with contact tracing, in years (M;IQR)** | 9 (4.8-14.8) | 11 (6-19.3) |

\*Consisting of Sint-Eustatius, Saba, and Bonaire
M: Median
IQR: Inter-quartile range

First, respondents were shown a webpage containing information and objectives of the study. Thereafter, they could give their informed consent before starting the questionnaire.

Second, respondents were shown a short video explaining online-RDD (displaying the PowerPoint presentation also used in Phase 1).

Third, respondents sequentially worked through each of the three hypothetical scenarios, where they responded to the developed statements. In each respective scenario, respondents were additionally asked if they would use online-RDD for CT if it were available at their respective PHS.

Fourth, at the end of the questionnaire, we asked for respondents’ general intention (outside the context of the hypothetical scenarios) to use online-RDD for CT.

The online questionnaire was distributed through the survey software Formdesk (https://en.formdesk.com) and took approximately 25 minutes to complete.

#### Statistical analyses

Descriptive analyses were conducted for respondents’ characteristics, for the statements in each scenario, and for intention to use online-RDD for CT (in each scenario and in general). Percentages were reported for all categorical variables (S2 Table 1). Distributions of continuous variables were checked using histograms and median and inter-quartile range (M;IQR) were reported.

For reporting purposes, we grouped respondents who agreed and respondents who very much agreed to all statements. The combined percentage of agreeing respondents was reported.

In order to check if general intention to use online-RDD for CT was associated with respondents’ characteristics, we first created a dichotomous intention variable. Respondents who very much agreed and agreed were grouped, as were respondents who were neutral, disagreed, or very much disagreed. We then checked associations using Chi-square tests. Fisher’s exact test was used when assumptions for the Chi-square test were violated (i.e. less than 80% of categories having an expected count of five or over).

The data used for analyses can be found in S3 Dataset. All analyses were conducted in Statistical Package for the Social Sciences (SPSS) v.24.

#### Ethical considerations

The Medical Ethical Committee of the University Medical Centre Utrecht exempted this study from medical scientific research requirements [reference number: 19-249/C].

## Results

### Study participants

Table 1 provides an overview of the characteristics of participants in the qualitative and the quantitative phases. We conducted twelve semi-structured interviews with PHPs; six (50%) with nurses and six (50%) with doctors. Interviewees had a median age of 38.5 years (IQR: 34.6-56.8) and a median of nine years (IQR: 4.8-14.8) of experience with CT. Four (33.3%) interviewees were male and eight (66.7%) were female.

We invited 260 Dutch PHS nurses and doctors to the online questionnaire. Of these, 81 started the online questionnaire (response rate: 31%). Ten respondents did not complete the online questionnaire. These were excluded from analyses. One respondent was also excluded for giving identical answers to all questions. The final number of included respondents was 70. Respondents had a median age of 49 years (36-59.3). Twenty-two respondents were male (31.4%) and 48 were female (68.6%). Most respondents were employed in the provinces Gelderland (20%) and North-Holland (18.6%). Thirty-three respondents (47.1%) were PHS-nurses, 35 (50%) were PHS-doctors, and two (2.9%) were department managers at a PHS. The latter two respondents were not excluded from analyses, since both reported having experience with CT. On average, respondents had a median of 11 years (IQR: 9-19.3) of experience with CT.

### Advantages and challenges of online-RDD for CT

Five themes related to advantages and challenges of online-RDD for CT were identified. Both qualitative and quantitative results are discussed in this section.

### Advantages of online-RDD for CT

Anticipated advantages of online-RDD for CT were related to: ‘accommodating easy and autonomous participation in CT for patients and contact persons’ and ‘reaching contact persons more efficiently in CT’. See Table 2 for illustrating quotes. See S2 Table 1 for a detailed overview of quantitative results.

**Table 2.** Quotes related to advantages of online-RDD for CT.

| Themes | Illustrative quotes |
| --- | --- |
| Accommodating easy and autonomous participation in CT for patients and contact persons. | "I think you can take away many barriers by having the index [patient] forward this [the online CT questionnaire]. Especially if it is possible to do so anonymously. For example, with scabies, all the bed-partners, and with mumps, all the kissing partners... We do not actually need to know all of that. They can just warn those themselves."<br>Nurse, mid-thirties |
|  | "In today's society, during the day people work, sleep, or are unavailable. This provides opportunities to go around that ... so those who are hard to reach by telephone could think "this is easy, I'll just do this tonight."<br>Nurse, mid-thirties |
| Reaching contact persons more efficiently in CT. | "I believe it's just more efficient to handle things this way [with online-RDD]. And if things can be done more efficiently, that appeals to me. It saves you time."<br>Doctor, late-twenties |
|  | "There is an advantage for the index [patient]. With the push of a button, he can just contact his whole group. And the information will come back quickly. So... I believe that is very efficient."<br>Doctor, mid-fifties |

#### Accommodating easy and autonomous participation in CT for patients and contact persons

Online-RDD was perceived by interviewees as a method that may accommodate patient and contact person participation in CT in different ways. First, direct patient-to-contact-person communication was perceived to lower barriers for patients who do not want to directly share (sensitive) information about their contact persons with PHPs. Second, the use of online CT-questionnaires, which may also be forwarded anonymously by patients to their contact persons, was considered an easy and low-threshold method for informing and/or warning contact persons. Third, online-RDD could give patients and contact persons the opportunity to participate in the CT-process at a moment of their choosing, rather than being dependent on contact with PHPs during office hours. Interviewees felt, however, that the aforementioned advantages may depend on the digital skills, literacy, and self-efficacy of patients and contact persons. In addition, it was anticipated that if information or warnings delivered in the CT-process would be experienced as particularly severe by patients and contact persons, this could inhibit their participation in CT through online-RDD.

A majority of questionnaire respondents believed that online-RDD would provide relatively easy and low-threshold options to inform and warn contact persons in all scenarios (94.3% scabies; 78.6% shigella; 52.9% mumps). Online-RDD was anticipated to be a more pleasant form of CT for patients and contact persons by 45.7% of questionnaire respondents in the scabies scenario, by 54.3% in the shigella scenario, and by 32.9% in the mumps scenario.

#### Reaching contact persons more efficiently in CT

Interviewees believed that with online-RDD, contact persons could be reached more efficiently in CT. For example, normally PHPs are tasked with gathering contact information on, and reaching out, to contact persons (often separately). If instead patients would self-identify their contact persons and send them a weblink to an online CT-questionnaire through ‘the push of a button’, this would save PHP’s time and labour. Consequently, it was felt that with online-RDD an increased number of contact persons could be reached in the CT-process, in a relatively short amount of time. This was especially considered advantageous if the scope of CT would be large, for example during outbreak investigations with many at-risk contact persons.

It was felt by 84.3% of questionnaire respondents that online-RDD could save time in CT in scabies scenario, compared to 70% in the shigella – and 50% in the mumps scenario. A lower workload through online-RDD was anticipated by 67.1%, 55.7%, and 37.2% of questionnaire respondents in the scabies, shigella, and mumps scenarios respectively. Compared to regular CT-methods, 61.4% (scabies), 55.7%(shigella), and 42.9% (mumps) of questionnaire respondents believed that they could reach more contact persons through online-RDD.

### Challenges for CT with online-RDD

Anticipated challenges for CT with online-RDD were related to: *limited opportunities for PHPs to support, motivate, and coordinate the execution of CT, adequately conveying measures to patients and contact persons, and anticipated unrest among patients and contact persons. See Table 3 for illustrating quotes*. See S2 Table 1 for a detailed overview of quantitative results.

**Table 3.** Quotes related to challenges for CT with online-RDD.

| Themes | Illustrative quotes |
| --- | --- |
| Limited opportunities for PHPs to support, motivate, and coordinate the execution of CT | "I do not know if you can really create a sense of urgency when you just send someone a web-link. Sometimes a PHS has a bit more authority, so that people really take it seriously."<br>Doctor, mid-fifties |
|  | "You let go of the part where you yourself call someone. The part of: 'will this be sent to the right people?, are we missing anyone?, are we not informing too many people?' You can try to incorporate that into the system, but that danger will always remain."<br>Nurse, early-forties |
| Adequately conveying measures to patients and contact persons | "Does someone understand what he is reading and what the consequences are? It makes you dependent of what the other person does. I do see it as an opportunity, but also as a risk to in the end not be able to execute the measures you would like to."<br>Nurse, early-thirties |
| Anticipated unrest among patients and contact persons | The feeling I get of people ... is that they appreciate to be talked to personally, so that we as professionals can explain why we call, and why we are asking questions. Then they can also ask their questions straight away. Then you can immediately take away a little bit of unrest. They immediately think the worst, that they are sick."<br>Nurse, early-forties |

#### Limited opportunities for PHPs to support, motivate, and coordinate the execution of CT

Interviewees felt that CT is a complex task in which, normally, PHPs hold a central role with regard to its execution. For example, PHPs may introduce the situation at hand to patients and their contact persons, support them where necessary, and motivate their cooperation. This, subsequently, allows PHPs to further identify - and carry out CT to -the right persons.

With online-RDD, interviewees worried that these elements of CT could potentially not be dealt with adequately. Contact between PHPs, patients, and contact persons would be reduced and information would mainly have to be communicated through an online CT-questionnaire. As such, patients and contact persons would operate relatively autonomously in identifying and reaching (relevant) contact persons. Though this was not inherently considered problematic, it was felt that with limited involvement of PHPs in the CT-process, patients and their contact persons might not take the CT-process seriously, not understand what is expected of them, or not want to cooperate.

Interviewees believed that these issues could lead to reaching irrelevant – or missing (information on) contact persons in the CT-process. Both these anticipated drawbacks could compromise the execution of CT and were considered particularly problematic if the consequences for individual or public health were potentially severe.

Compared to regular-CT methods, online-RDD was felt to limit PHP-involvement in CT by 58.6% of questionnaire respondents in the scabies scenario, compared to 42.9% in the shigella scenario and 50% in the mumps scenario. With online-RDD, 27.2% (scabies), 20% (shigella) and 50% (mumps) of questionnaire respondents anticipated that they could not adequately support patients and contact persons in the CT-process. It was believed by 37.1%, 52.9%, and 32.9% of questionnaire respondents, in the scabies, shigella, and mumps scenarios respectively, that they could miss contact persons through online-RDD.

#### Adequately conveying measures to patients and contact persons

With limited involvement of PHPs in the CT-process and information being communicated mainly through an online CT-questionnaire, interviewees additionally worried about adequately communicating, and subsequently delivering, recommended or required measures (e.g. instructions to seek treatment, maintain a hygienic routine, isolate/quarantine, etc.) to patients and contact persons. If done inadequately, it was anticipated that patients and their contact persons may, willingly or unwillingly, not (sufficiently) undertake the necessary precautions or actions. This was considered especially challenging if available measures were relatively complex or impactful.

Compared to regular CT-methods, 50% (scabies), 38.6% (shigella), and 51.4% (mumps) of questionnaire respondents anticipated that they could less adequately convey measures to patients and contact persons through online-RDD.

#### Anticipated unrest among patients and contact persons

Interviewees were concerned that patients and contact persons may become overly worried when receiving a notification of potential exposure to an infectious disease, especially without introduction or support from a PHP, which could compromise the execution of CT. This was considered particularly challenging if the consequences of a particular disease could be experienced as severe, or sensitive (stigmatised) by patients and contact persons. In addition, concerns were expressed in regard to people excessively sharing the online CT-questionnaire, which could lead to unrest ‘spreading’ through online-RDD.

Compared to regular CT-methods, 74.3% (scabies), 25.7% (shigella), and 62.9% (mumps) of questionnaire respondents anticipated that online-RDD could lead to unnecessary unrest among patients and contact persons.

### Intention to use online-RDD for CT

Overall, interviewees indicated that they would want to use online-RDD for CT, as they considered it a useful, addition tool to regular CT-methods. A majority of questionnaire respondents (71%) similarly indicated that they would want to use online-RDD for CT. None of respondents’ characteristics were associated with intention to use online-RDD (S2 Table 2)

Nevertheless, interviewees indicated that the potential application of online-RDD in practice depends on various circumstances (as outlined earlier). This was similarly reflected by questionnaire respondents, of whom 77.1% and 61.4% indicated that they would want to use online-RDD for CT in the scabies and the shigella scenarios, compared to 37.1% in the mumps scenario.

## Discussion

### Principal findings

This is the first study that investigates how Dutch PHPs involved in CT perceive the potential application of online-RDD for CT of infectious diseases that spread through close contact between individuals. Online-RDD was anticipated to benefit public health practice, as indicated by 71% of questionnaire respondents with a favourable intention towards using online-RDD for CT in general. PHPs anticipated that online-RDD could enhance CT through accommodating easy and autonomous participation in CT for patients and their contact persons, and reaching contact persons more efficiently in CT. Challenges were anticipated in regard to limited opportunities for PHPs to support, motivate, and coordinate the execution of CT, adequately conveying measures to patients and contact persons, and anticipated unrest among patients and contact persons. PHPs considered online-RDD a suitable addition (rather than alternative) to regular CT-methods, depending on circumstances under which CT is applied. Online-RDD was considered most applicable for CT when it was anticipated that patients and contact persons are reluctant to share (sensitive) information directly with PHPs, digitally skilled and literate individuals are involved, and the scope of CT is large (many contact persons need to be reached). Online-RDD was considered less applicable when consequences of missing information or individuals in CT are potentially severe for individuals or public health, when measures that patients and contact persons need to undertake are relatively complex or impactful, and when a disease is perceived as particularly severe or sensitive by patients and their contact persons.

PHPs’ intention to use online-RDD for CT differed strongly between the hypothetical scenarios used in this research (scabies 77.1%; shigella 61.4%; mumps 37.1%). The extent to which online-RDD’s advantages and challenges for CT were anticipated by questionnaire respondents similarly differed between the scenarios: advantages were most frequently anticipated in the scabies and the shigella scenarios; disadvantages most frequently in the mumps scenario. These findings underscore the importance of the previously outlined characteristics of situations in which online-RDD may be applied for CT. For example, questionnaire respondents (much like interviewees) may have considered the scabies scenario particularly suited for CT due to the involvement of students (who typically considered as relatively digitally skilled and literate), and the likely involvement of many contact persons (due to the index being described as living in a ‘large scale student housing complex’). In comparison, the mumps scenario gives no reason to believe that many contact persons are involved, but involves an exposed infant, which may demand action from contact persons (e.g. post-exposure prophylaxis) and comprises a potentially sensitive situation. In summary, it is clear that online-RDD may be particularly useful to public health practice as an additional (complementary) method for CT, rather than as an alternative to regular CT.

Online-RDD’s anticipated advantages and challenges for CT are, to a large extent, related to less involvement of PHPs, and conversely, a more autonomous role of patients and contact persons in the CT-process. In a broad sense, these are commonly described topics in eHealth implementation studies (19). However, literature specifically related to PHPs’ perception on the implementation of (online) innovations in CT is scarce. One closely related subject is (online) partner notification in the context of sexually transmitted diseases, which resembles online-RDD in the sense that patients inform/warn their contact persons. Two studies that investigated healthcare providers’ perspectives of partner notification yielded similar results to those presented in this study: PHPs believed that they could reach more contact persons through partner notification on the one hand, but had concerns regarding patients’ commitment to reach out to their contact persons on the other hand (20,21). These similarities indicate that strategies used to overcome challenges in the field of sexually transmitted diseases, such as motivational interviewing with index patients, may be similarly useful with online-RDD.

### Strengths and limitations

One important strength of this research was the mixed methods design, which allowed us to study qualitative findings in a larger population of PHPs (15). We also developed and extensively pilot tested our research materials, in particular the audio-visual introduction to online-RDD and the hypothetical scenarios, in close collaboration with PHPs. This allowed respondents to thoroughly deliberate on the use of online-RDD for CT, despite the conceptual nature of this exercise.

However, a number of limitations need to be addressed. First, at the time of this study online-RDD was unknown to the majority of Dutch PHPs. Therefore, it should be taken into account that the information provided to research participants through this study shaped, at least to some extent, participants’ expectations of online-RDD for CT in practice. Second, this research was conducted just before the COVID-19 outbreak. Considering the impact of the outbreak and the increasing interest in - and debate on technological innovations in CT, we are unsure to what degree this may have affected PHPs perceptions of online-RDD as presented in this study. Third, the limited sample size of this research’s quantitative component prohibited multi-variable or other explanatory statistical analyses. Nevertheless, considering the questionnaire’s response rate (31%) and geographical coverage (all provinces in the Netherlands), we believe that our results still give useful insights in Dutch PHPs’ perception of online-RDD for CT.

### Online-RDD and COVID-19

Based on this study, and based on our previous work conducted with online-RDD (12,14,22), we believe that online-RDD may be particularly useful as an additional method to regular CT in the context of COVID-19. PHPs anticipated that online-RDD could save time in CT, reach more contact persons, and decrease PHPs’ workload, which are highly valuable assets considering the unprecedented scope of the COVID-19 pandemic and the rapid spread of the virus. Nevertheless, considering the potential severity of COVID-19 for individuals and public health, the anticipated challenges of COVID-19 (e.g. missing contact persons in CT, potential unrest among patients and contact persons) warrant attention too.

In order to apply and investigate online-RDD’s anticipated advantages and challenges for CT during the ongoing COVID-19 pandemic, we propose a staggered implementation study (see Fig 2). This entails working towards ‘full’ online-RDD in a series of stages, in which proceeding to a next stage is dependent on its success in earlier stages. Such an incremental design is known to be effective in the context of eHealth implementation (19), and leaves room to sufficiently address the challenges of online-RDD anticipated by PHPs.

**Fig 2.**
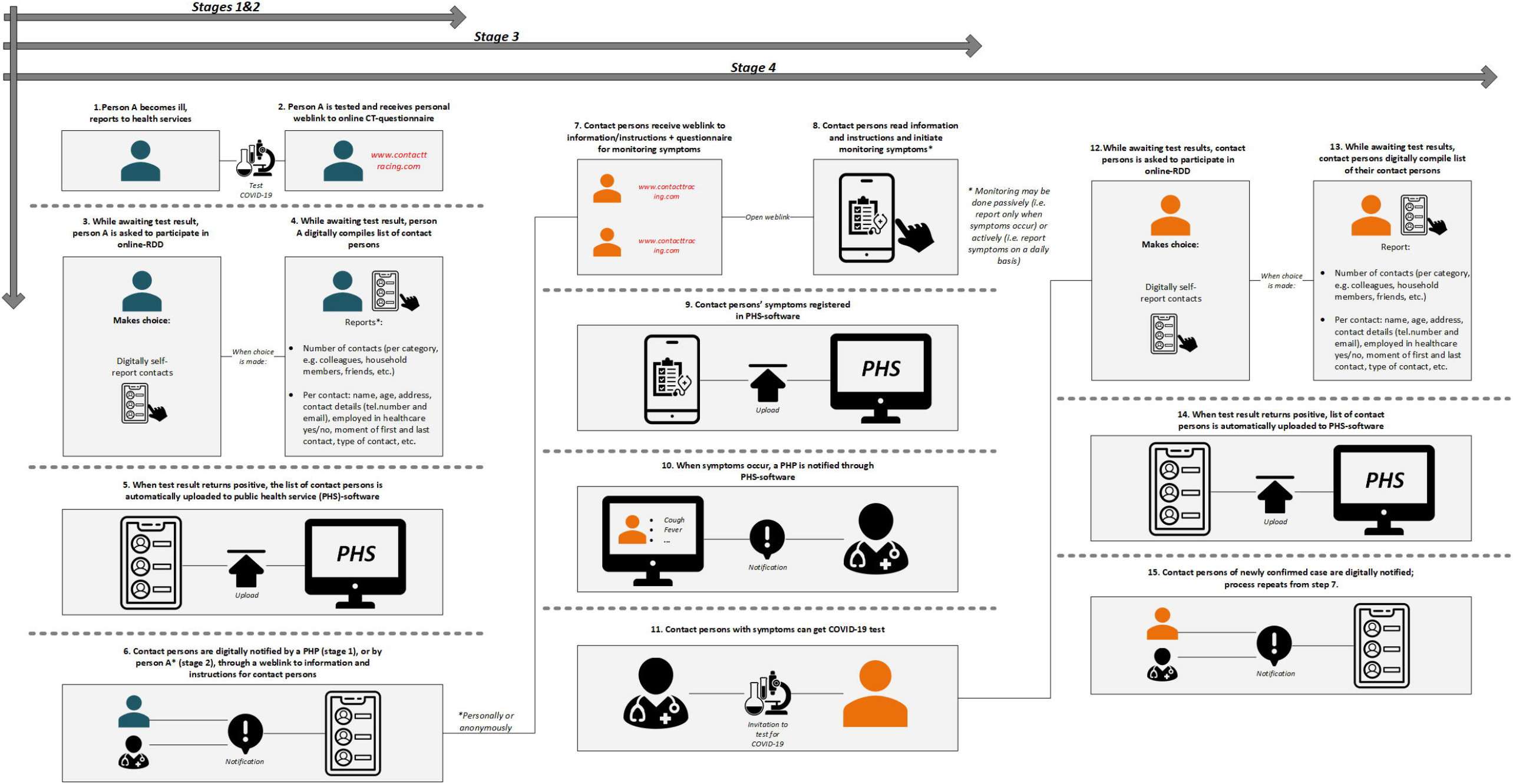
Online-RDD staggered implementation study design.

In a first stage, PHPs could provide patients with online guidelines for identifying relevant contact persons and to ask patients to digitally self-report their contact persons, and to provide information about these persons. PHPs could then review the reported list of contact persons and focus CT on those contact persons they consider relevant. In a second stage, PHPs may or may not review the contact list provided, after which patients themselves can, personally or anonymously, contact their contact persons and digitally send them information and instructions that would normally be conveyed by PHPs over the phone or via email. In a third stage, contact persons who received a notification are asked to digitally keep track of and report their symptoms. If symptoms present, PHS are notified, and a PHP can get in touch with the respective contact person(s). In a fourth stage, contact persons keep track of and report their symptoms. If symptoms present, contact persons are directly prompted to self-report their own contact persons. If a contact person tests positive for COVID-19, the contact list is readily available and a PHP, or the newly diagnosed patient may immediately reach out to his/her contact persons.

During (and after) this implementation study, we believe that it will be necessary to let patients and contact persons choose, at all times, whether they would like to participate in online-RDD or in regular CT. Some patients will feel more or less comfortable in using online-RDD, and also contact persons may not feel as comfortable with online-RDD as patients sending them the questionnaire, instead of a PHP. In addition, there should be opportunities for contact persons of patients to reach out to PHPs for clarification or support during the CT process and vice versa. Therefore, it seems beneficial to the CT process to facilitate a flexible integration of on- and offline components in the CT process.

In addition, it will be of crucial importance to also investigate the (acceptability of the) potential implementation of online-RDD from the perspective of (potential) patients and contact persons. This may be investigated in the context of the above outlined implementation study, but should rather be investigated in a similar fashion to the present study.

## Conclusion

In this study, we have shown that from PHPs’ perspective, online-RDD may be a welcome, additional method for CT. Advantages of online-RDD for CT were anticipated in regard to accommodating easy and autonomous participation in CT for patients and contact persons, and reaching contact persons more efficiently during CT. Challenges were anticipated in regard to limited opportunities for PHPs to support, motivate, and coordinate the execution of CT, adequately conveying measures to patients and contact persons, and anticipated unrest among patients and contact persons. We propose a staggered implementation study to further investigate the use of online-RDD for enhanced CT during the current COVID-19 pandemic.

## Data Availability

Yes - all data are fully available without restriction

## Supporting information

**S1 Figs. Scabies, shigella & mumps hypothetical scenario’s**.

**S2 Tables. 1) Questionnaire respondents’ responses to statements based on qualitatively identified themes; 2) Associations between questionnaire respondents’ characteristics and intention to use online-RDD for CT**.

**S3 Dataset. Quantitative data used for analysis**

